# Effects of different exercise training program on post-exercise V̇O_2_ kinetics and V̇O_2_ recovery delay in stable patients with coronary heart disease

**DOI:** 10.1101/2025.05.09.25325944

**Authors:** Mathieu Gayda, Lukas-Daniel Trachsel, Pierre-Marie Leprêtre, Florent Besnier, Maxime Boidin, Julie Lalongé, Louis Bherer, Martin Juneau, Anil Nigam

## Abstract

Post-exercise V̇O_2_ kinetics and V̇O_2_ recovery delay (RD) are clinical prognostic markers in cardiac patients, but have not been studied after exercise training in patients with coronary heart disease (CHD). We aimed to compare the effects of 12-weeks moderate-intensity continuous exercise training (MICET), low volume high-intensity interval training (LV-HIIT), or combined MICET/HIIT on O_2_ deficit, post-exercise V̇O_2_ kinetics, O_2_ debt and V̇O_2_ recovery delay (RD) in patients with CHD.

**Methods:** Patients with CHD were randomised in MICET, LV-HIIT or combined MICET/HIIT group for 12 weeks. Cardiopulmonary exercise test (CPET) parameters were assessed, and key exercise variables were calculated during and after exercise. CPET post-exercise kinetics time constant (r) (for V̇O_2_, V̇CO_2_, *V̇*E and HR), O_2_ deficit, O_2_ debt and V̇O_2_ recovery delay (RD) were calculated before and after training.

**Results:** A significant time effect (training) for r V̇O_2_ (min) (p<0.05) was shown for all groups. Shorter r V̇O_2_ values with small effect size (ES: 0.21 to 0.4) were noted for the combined MICET/HIIT and MICET groups. A significant time effect (p<0.01) was noted for O_2_ debt that was increased after training (ES: 0.1 to 0.47). No significant statistical effect was shown for V̇O_2_ RD and r V̇CO_2_, r *V̇*E, r HR and O_2_ deficit in all groups.

**Conclusions:** In patients with CHD, exercise training improved post-exercise V̇O_2_ kinetic and the O_2_ debt, with a higher impact of exercise dose (combined MICET/HIIT). Exercise training did not improved the V̇O_2_ RD or other *τ* CPET recovery variables in CHD patients.

## Introduction

Cardiopulmonary exercise testing (CPET) is the gold standard to assess cardiorespiratory fitness (*V̇*O_2_peak), a powerful indicator of disease severity and prognosis in patients with coronary heart disease (CHD) ^1, 2^. Exercise-based secondary prevention programs can improve *V̇O*_2_peak that is associated with a reduction of cardiovascular mortality and events in patients with CHD ^3–5^. In addition to *V̇*O_2_peak, other cardiopulmonary variables measured during CPET and its early post-exercise recovery phase can provide important complementary physiologic and metabolic information after exercise exposure of cardiac patients. Post-exercise *V̇*O_2_ kinetics is partly related to the recovery of phosphocreatine supply in the muscle, and is very sensitive to impairment of the O_2_ delivery and diffusion in patients with chronic heart failure (CHF) ^6–8^. *V̇*O_2_ kinetics during the early post-exercise recovery phase are considered independent of the exercise level but these values are affected by severe reduction in exercise capacity and abnormal cardiac response to exercise ^9, 10^. After exercise cessation, prolonged *V̇*O_2_, ventilation (*V̇*E) and heart rate recovery kinetics has been associated with a worse clinical profile and outcomes in patients with CHF ^6, 8, 11^. *V̇*O_2_ kinetics and O_2_ debt, i.e. the difference between the resting rate of oxygen consumption and the elevated rate following an exercise, are also affected by the hemodynamic negative effect of not providing the body’s requested amount of O_2_ generated during exercise (O_2_ deficit) with values correlated with O_2_ exercise delivery and exercise tolerance ^10^. The value of the peak oxygen recovery delay (V̇O_2_ RD), as defined as the time from the beginning of recovery until the V̇O_2_ permanently falls below V̇O_2_ peak, was also inversly related to cardiac output reserve, predicts time to heart transplantation and was associated with disease severity in patients with CHF ^6, 11^. In patients with CHD, abnormalities in post-exercise *V̇*O_2_ kinetics shape («Hump») and slowed *V̇*O_2_ kinetics have been related to myocardial ischemia ^12, 13^, while time constant (r) of V̇O_2_, V̇CO_2_ and heart rate were not different between healthy subjects and patients with CHD ^10^. With these conflicting results, the prevalence of the V̇O_2_ RD and its hemodynamic relationship has not been studied in patients with CHD. Previous study in healthy subjects suggested that initial aerobic fitness and exercise training improved post exercise *V̇*O_2_ kinetics ^14–16^. In patients with CHF, previous studies demonstrated that various exercise training program (aerobic, resistance plus respiratory or interval training) improved post-exercise *V̇*O_2_ kinetics after submaximal ^17, 18^ or maximal exercise ^19^, via mainly improvement of muscular O_2_ delivery. There are actually no studies investigating the effects of different exercise training programs on O_2_ deficit, post-exercise *V̇*O_2_ kinetics, O_2_ debt, and the V̇O_2_ RD in patients with CHD. The main aims of our study were to assess the effects of moderate-intensity continuous exercise training (MICET), low volume high-intensity interval training (LV-HIIT), or combined MICET/HIIT on O_2_ deficit, post-exercise V̇O_2_ kinetics, O_2_ debt, and V̇O_2_ RD in patients with CHD. We hypothesised that exercise training will improve post-exercise *V̇*O_2_ kinetics and the V̇O_2_ RD in patients with CHD, with stronger effects for higher exercise dose.

## Material and methods

### Study design and participants

This retrospective study is based on data from three prospective randomized exercise intervention trials conducted in patients with CHD ^20–24^. All studies protocols were approved by the Research Ethics and New Technology Development Committee of the Montreal Heart Institute, and were registered on ClinicalTrials.gov (ClinicalTrials.gov identifier numbers: NCT03414996, NCT02048696, NCT03443193) and written informed consent was obtained by each patient. All patients were referred for a structured aerobic exercise training program at the ÉPIC Center of the Montreal Heart Institute and were involved in a training intervention study. All patients with CHD were under optimal medical therapy after coronary revascularisation.

### Measurement

Baseline clinical assessment (i.e. medical history, physical examination and anthropometric measurements, including body composition analysis (bioimpedance, Tanita, model BC418, Japan), transthoracic echocardiography (GE, Vivid 9, USA) and CPET were performed at baseline and after completion of each of the program.

### Maximal cardiopulmonary exercise testing (CPET)

Maximal CPET was performed on a cycle ergometer (Ergoline 800S, Bitz, Germany) according to the latest join recommendations and as previously published ^21, 22, 24, 25^. Following a 3-minute warm up phase at an initial work load of 20 W, an incremental exercise test with 15 Watt increments per min until exhaustion at a pedalling speed > 60 rpm was performed. The recovery phase consisted of 2 minutes of active recovery at 20 W at pedalling speed between 50 and 60 rpm, followed by 3 minutes of passive recovery. Gas exchange parameters were continuously measured at rest, during exercise, and during recovery using a metabolic system (Oxycon Pro, CareFusion, Jaeger, Germany), breath by breath basis and then averaged every 15 sec. as recently published ^21–24^. There was a continuous ECG monitoring (Marquette, case 12, St. Louis, Missouri). Blood pressure and rate of perceived exertion (RPE) were measured each 3 min throughout the test. The highest V̇O_2_ value reached during the exercise phase was considered as the V̇O_2_ peak, and this value was used for the calculation of the O_2_ pulse (V̇O_2_ /HR) according to the recent recommendations ^21, 22, 24, 25^.

### Oxygen deficit calculation

O_2_ deficit was calculated by subtracting the measured V̇O_2_ during exercise phase (warm-up and incremental exercise phase) from the theoretical V̇O_2_ value computed based on the energy cost to develop 1 watt on an electromagnetic brake ergometer ^26^.

### Post exercise CPET variables

Recovery (or post-exercise) was defined by the period after exercise cessation at peak effort and was activated manually by the operator during CPET testing. Recovery CPET variable were measured during 5 minutes (2 min of active recovery and then 3 min of passive recovery). The post exercise CPET kinetics (V̇O_2_, V̇CO_2_, *V̇*E) were assessed with Oxycon Pro software utilities Tau (*τ)* mathematical function (Oxycon Pro CareFusion, Jaeger, Germany), indicated by mono exponential function ^8, 27, 28^ :

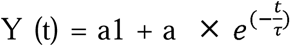 where a1 is the CPET (V̇O_2_, V̇CO_2_, *V̇*E, HR) value at the beginning of recovery, a is the asymptote value (value at end of 5 min-recovery), r is tau (or time constant : time need to reach 63% of the gain) ^8, 27, 28^. The V̇O_2_ recovery delay (V̇O_2_ RD) was calculated according to recent previous methodology ^6^ and defined as the time from the beginning of recovery until the V̇O_2_ permanently falls below V̇O_2_ peak. As well, heart rate recovery (HRR) at 1 minute was calculated by subtracting heart rate at 1 min recovery from the maximal heart rate at peak exercise ^20^. O_2_ debt was calculated by taking the exercise-final V̇O_2_ as the peak of the curve and integrating V̇O_2_ from there until it decayed to the rest value ^26^.

### Exercise training intervention (aerobic and resistance training)

The aerobic exercise training intervention consisted of three different training modalities: moderate-intensity continuous exercise training (MICET), low volume high-intensity interval training (LV-HIIT), and a combined HIIT/MICET, as previously reported ^20–24^. All patients performed 2 to 3 exercise training sessions a week on a bicycle ergometer. Resistance training (RT) was then performed following each endurance session ^21, 22, 24^. The training load was calculated for the aerobic training component according to the adapted model of Calvert et al. ^29^. More details regarding aerobic and resistance training protocols are available **in Supplementary materials File S1.**

### Statistical analyses

Data are presented as means ± standard deviation for continuous variables and presented as frequencies and percentages for categorical variables. Statistical analyses were performed using GraphPad Prism 9.0 (GraphPad Software, Inc., La Jolla, California, USA). Normal distribution was verified with a Shapiro-Wilk test and baseline characteristics were compared between groups using either a *t*-test or a Mann-Whitney test, a chi-square test was used for categorical variables. Mixed model analysis (groups x time) was used to study the CPET parameters across time and between groups. Models with group, time and group x time interaction as independent variables were used. The group x time interaction was the main focus of the analysis as it tested the difference in the change (post-pre) between the three groups. Multiple comparison test was done using Šídák test to localize differences, with adjusted p-value. The magnitude of the effect size was compared using Cohen’s d scale and was considered either trivial (d <0.2), small (0.2 < d < 0.5), moderate (0.5 < d < 0.8) or large (d > 0.8). A *p*-value < 0.05 was considered statistically significant.

## Results

### Clinical characteristics

The clinical characteristics are presented in table 1. In the final analysis, we included a total of 82 patients with CHD (Combined MICET/HIIT=38, HIIT n=26, MICET=18). Patients in the combined MICET/HIIT had a lower prevalence of post ACS, lower use of dual antiplatelet therapy, higher weekly training sessions (p<0.05) vs. the other groups (HIIT, MICET). Resting DBP was higher for combined HIIT/HIIT vs. HIIT (p<0.05). Resting LVEF was lower in the MICET group vs. the 2 others (p<0.05). Training load was higher for Combined MICET/HIIT and MICET vs. HIIT (p<0.0001). Otherwise, there were no differences with regards to baseline clinical characteristics **(Table 1)**.

**Table 1:**
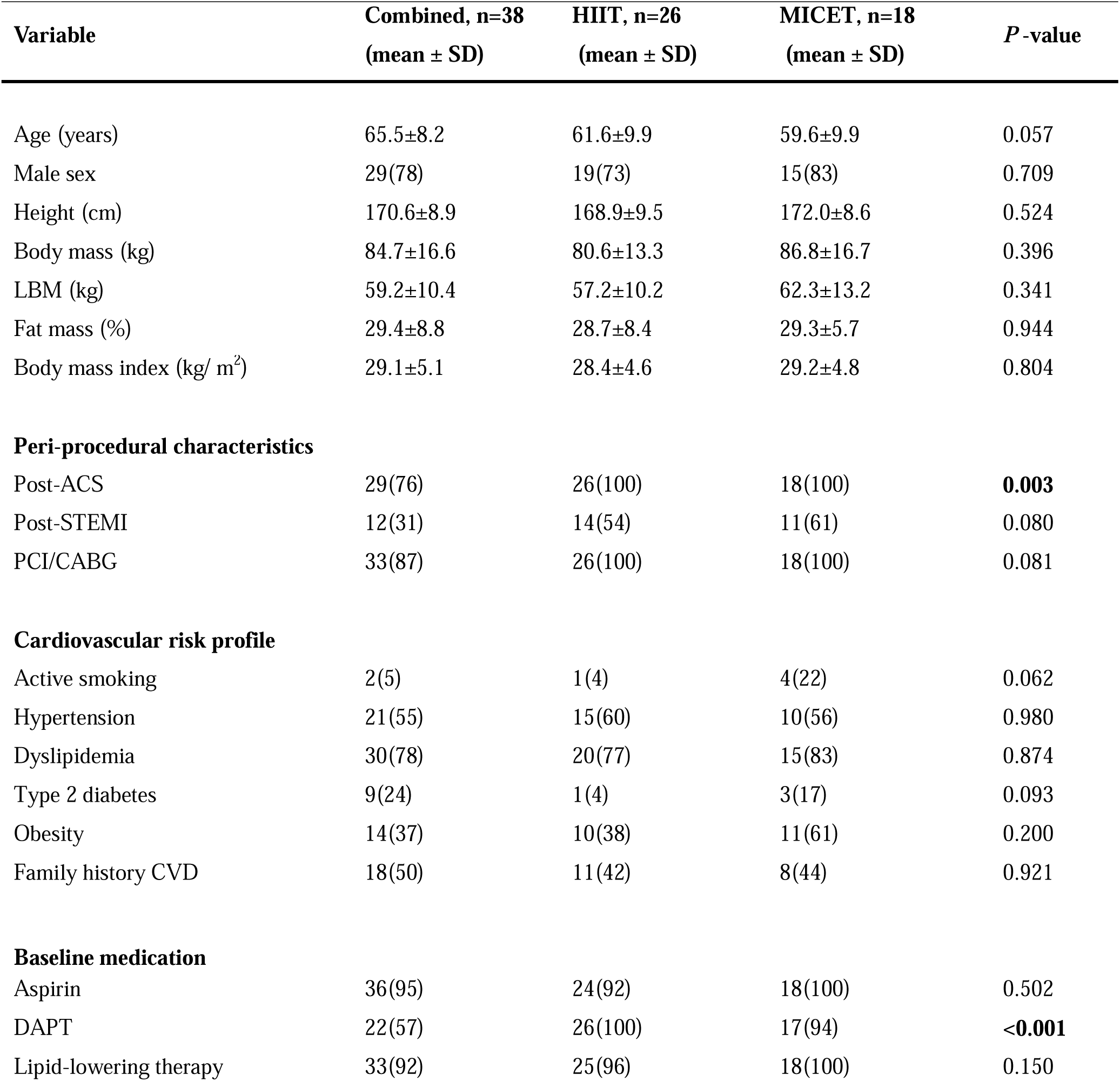
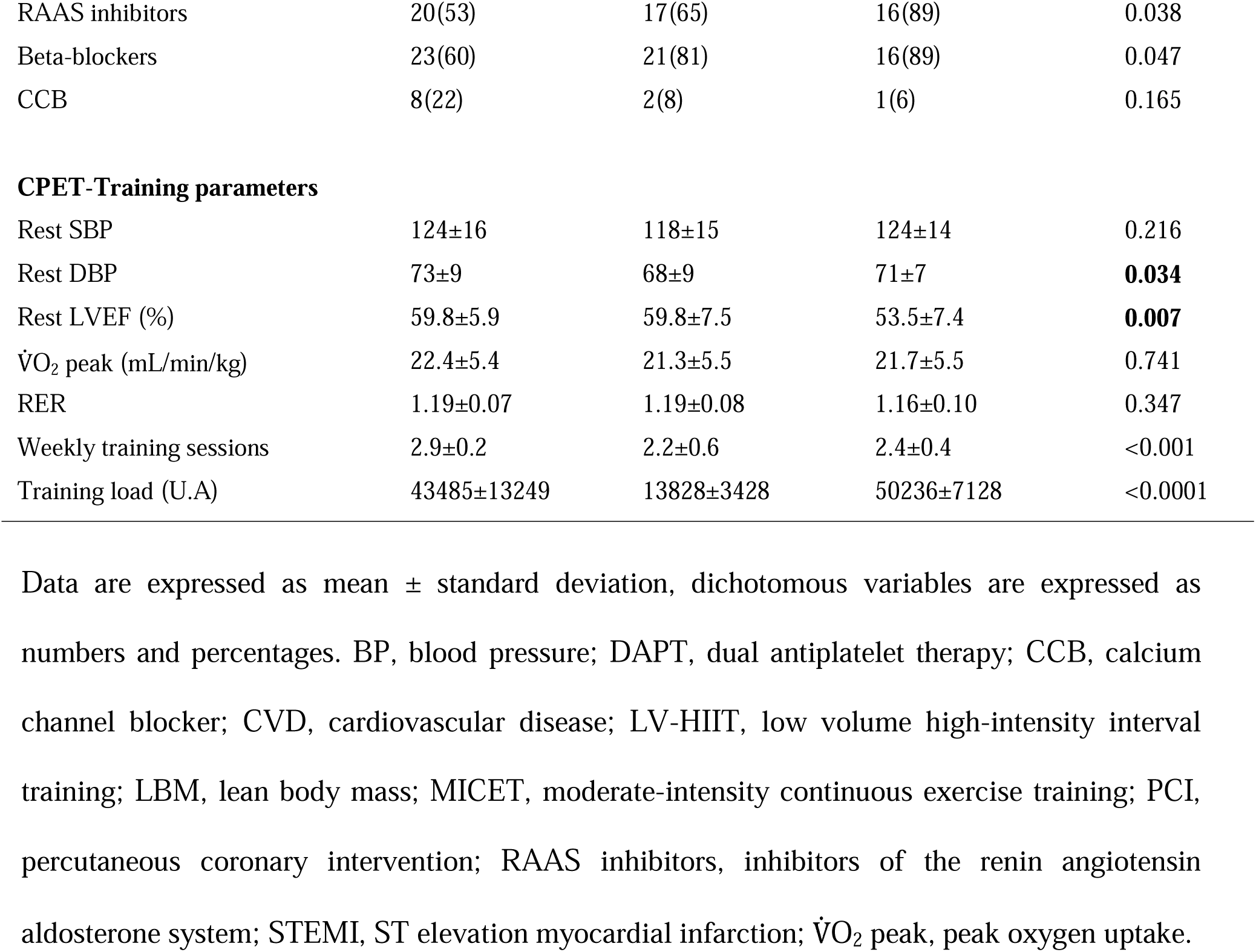
Clinical characteristics of patients with CHD randomized to combined MICET/HIIT, HIIT, or MICET.

| Variable | Combined, n=38<br>(mean ± SD) | HIIT, n=26<br>(mean ± SD) | MICET, n=18<br>(mean ± SD) | *P* -value |
| --- | --- | --- | --- | --- |
| Age (years) | 65.5±8.2 | 61.6±9.9 | 59.6±9.9 | 0.057 |
| Male sex | 29(78) | 19(73) | 15(83) | 0.709 |
| Height (cm) | 170.6±8.9 | 168.9±9.5 | 172.0±8.6 | 0.524 |
| Body mass (kg) | 84.7±16.6 | 80.6±13.3 | 86.8±16.7 | 0.396 |
| LBM (kg) | 59.2±10.4 | 57.2±10.2 | 62.3±13.2 | 0.341 |
| Fat mass (%) | 29.4±8.8 | 28.7±8.4 | 29.3±5.7 | 0.944 |
| Body mass index (kg/ m<sup>2</sup>) | 29.1±5.1 | 28.4±4.6 | 29.2±4.8 | 0.804 |
| **Peri-procedural characteristics** |  |  |  |  |
| Post-ACS | 29(76) | 26(100) | 18(100) | **0.003** |
| Post-STEMI | 12(31) | 14(54) | 11(61) | 0.080 |
| PCI/CABG | 33(87) | 26(100) | 18(100) | 0.081 |
| **Cardiovascular risk profile** |  |  |  |  |
| Active smoking | 2(5) | 1(4) | 4(22) | 0.062 |
| Hypertension | 21(55) | 15(60) | 10(56) | 0.980 |
| Dyslipidemia | 30(78) | 20(77) | 15(83) | 0.874 |
| Type 2 diabetes | 9(24) | 1(4) | 3(17) | 0.093 |
| Obesity | 14(37) | 10(38) | 11(61) | 0.200 |
| Family history CVD | 18(50) | 11(42) | 8(44) | 0.921 |
| **Baseline medication** |  |  |  |  |
| Aspirin | 36(95) | 24(92) | 18(100) | 0.502 |
| DAPT | 22(57) | 26(100) | 17(94) | **<0.001** |
| Lipid-lowering therapy | 33(92) | 25(96) | 18(100) | 0.150 |
| RAAS inhibitors | 20(53) | 17(65) | 16(89) | 0.038 |
| Beta-blockers | 23(60) | 21(81) | 16(89) | 0.047 |
| CCB | 8(22) | 2(8) | 1(6) | 0.165 |
| CPET-Training parameters |  |  |  |  |
| Rest SBP | 124±16 | 118±15 | 124±14 | 0.216 |
| Rest DBP | 73±9 | 68±9 | 71±7 | **0.034** |
| Rest LVEF (%) | 59.8±5.9 | 59.8±7.5 | 53.5±7.4 | **0.007** |
| $\dot{V}O_2$ peak (mL/min/kg) | 22.4±5.4 | 21.3±5.5 | 21.7±5.5 | 0.741 |
| RER | 1.19±0.07 | 1.19±0.08 | 1.16±0.10 | 0.347 |
| Weekly training sessions | 2.9±0.2 | 2.2±0.6 | 2.4±0.4 | <0.001 |
| Training load (U.A) | 43485±13249 | 13828±3428 | 50236±7128 | <0.0001 |

Data are expressed as mean ± standard deviation, dichotomous variables are expressed as numbers and percentages. BP, blood pressure; DAPT, dual antiplatelet therapy; CCB, calcium channel blocker; CVD, cardiovascular disease; LV-HIIT, low volume high-intensity interval training; LBM, lean body mass; MICET, moderate-intensity continuous exercise training; PCI, percutaneous coronary intervention; RAAS inhibitors, inhibitors of the renin angiotensin aldosterone system; STEMI, ST elevation myocardial infarction; $\dot{V}O_2$ peak, peak oxygen uptake.

### Exercise and recovery CPET parameters

The exercise and recovery CPET parameters for the three groups (combined MICET-HIIT, HIIT, MICET) are presented **in table 2**. There was significant time effect (training) regarding *τ* V̇O_2_ (min) (p<0.01) with no interaction. The *τ* V̇O_2_ was shorter with trivial to small effect size for the combined MICET-HIIT and MICET groups (ES: 0.21 and 0.40). No significant time, group or interaction effect was noted for *τ* V̇CO_2_, *τ V̇*E, *τ* HR, V̇O_2_ RD and O_2_ deficit for all groups (p>0.05). The prevalence of V̇O_2_ RD was of 43% (pre: n=35) at baseline and of 47% after training (n=38), and remained unchanged (p=0.63). A significant time effect (p<0.01) was shown for the O_2_ debt that was increased after training, with trivial to small effect size (ES: 0.10 to 0.47). There was no significant time, group or interaction effect for HRR at 1 min (p>0.05). A significant interaction and time effect (p<0.05) was found for O_2_ pulse and values were improved after training in the combined HIIT/MICET, HIIT groups and particularly in the MICET group (ES: 0.27 to 0.59). The training load was lower in the LV-HIIT group vs the 2 others groups (P<0.0001) and higher in the MICET group vs. combined HIIT/MICET (P<0.05) **(see Table 1)**.

**Table 2:**
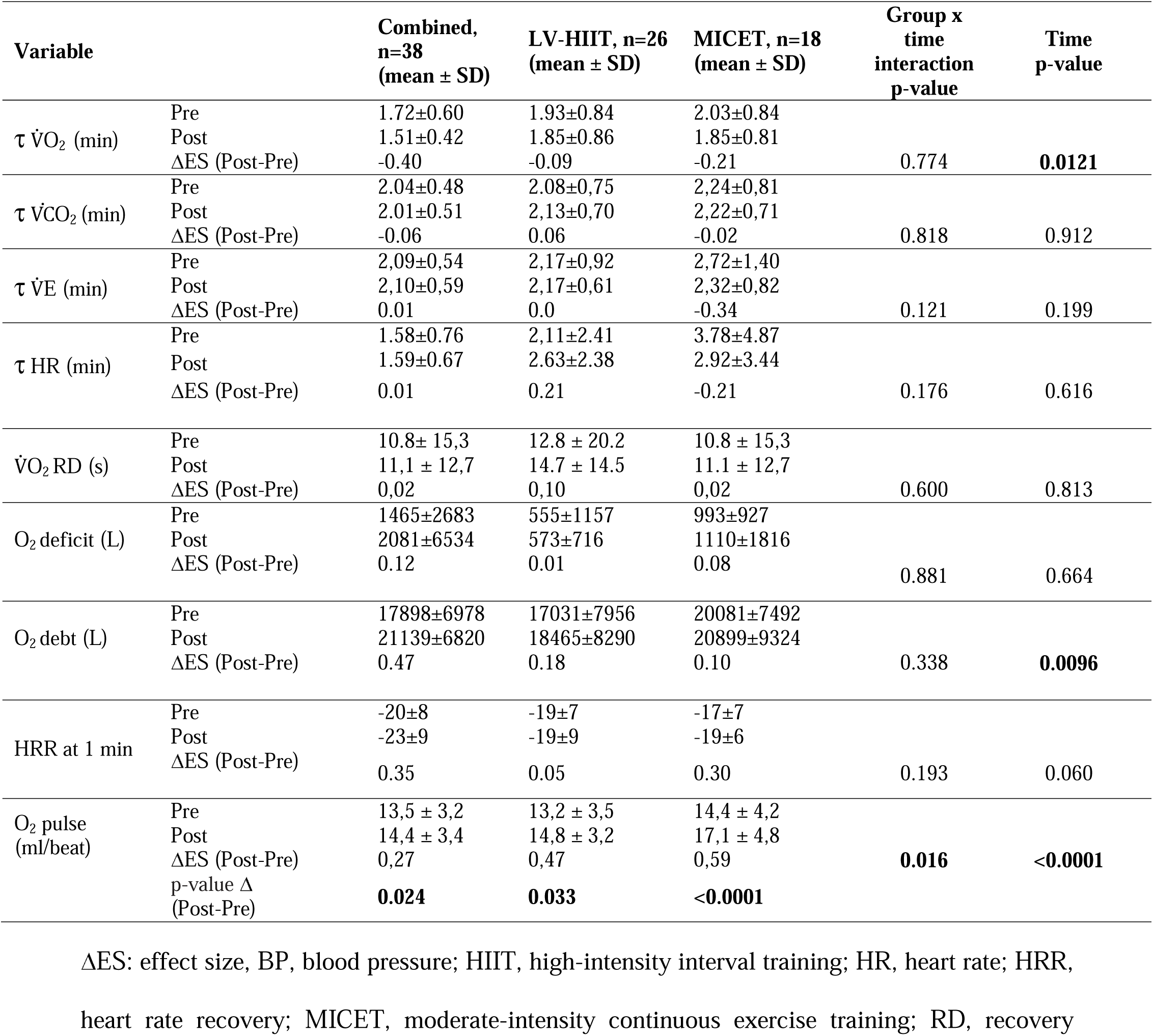
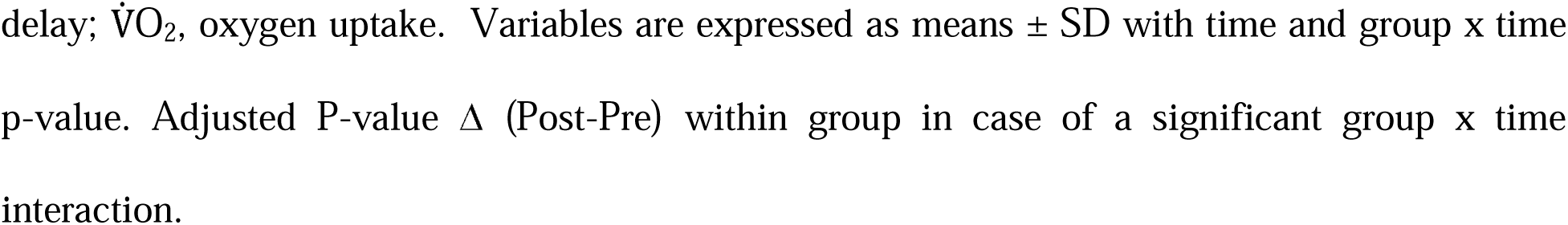
Exercise and recovery cardiopulmonary exercise test parameters in CHD patients randomized to combined MICET/HIIT, LV-HIIT, or MICET.

| Variable |  | Combined,<br>n=38<br>(mean ± SD) | LV-HIIT, n=26<br>(mean ± SD) | MICET, n=18<br>(mean ± SD) | Group x<br>time<br>interaction<br>p-value | Time<br>p-value |
| --- | --- | --- | --- | --- | --- | --- |
| $\tau \dot{V}O_2$ (min) | Pre | 1.72±0.60 | 1.93±0.84 | 2.03±0.84 | 0.774 | **0.0121** |
|  | Post | 1.51±0.42 | 1.85±0.86 | 1.85±0.81 |  |  |
|  | ΔES (Post-Pre) | -0.40 | -0.09 | -0.21 |  |  |
| $\tau \dot{V}CO_2$ (min) | Pre | 2.04±0.48 | 2.08±0.75 | 2.24±0.81 | 0.818 | 0.912 |
|  | Post | 2.01±0.51 | 2.13±0.70 | 2.22±0.71 |  |  |
|  | ΔES (Post-Pre) | -0.06 | 0.06 | -0.02 |  |  |
| $\tau \dot{V}E$ (min) | Pre | 2.09±0.54 | 2.17±0.92 | 2.72±1.40 | 0.121 | 0.199 |
|  | Post | 2.10±0.59 | 2.17±0.61 | 2.32±0.82 |  |  |
|  | ΔES (Post-Pre) | 0.01 | 0.0 | -0.34 |  |  |
| $\tau$ HR (min) | Pre | 1.58±0.76 | 2.11±2.41 | 3.78±4.87 | 0.176 | 0.616 |
|  | Post | 1.59±0.67 | 2.63±2.38 | 2.92±3.44 |  |  |
|  | ΔES (Post-Pre) | 0.01 | 0.21 | -0.21 |  |  |
| $\dot{V}O_2$ RD (s) | Pre | 10.8± 15,3 | 12.8 ± 20.2 | 10.8 ± 15,3 | 0.600 | 0.813 |
|  | Post | 11,1 ± 12,7 | 14.7 ± 14.5 | 11.1 ± 12,7 |  |  |
|  | ΔES (Post-Pre) | 0,02 | 0,10 | 0,02 |  |  |
| O<sub>2</sub> deficit (L) | Pre | 1465±2683 | 555±1157 | 993±927 | 0.881 | 0.664 |
|  | Post | 2081±6534 | 573±716 | 1110±1816 |  |  |
|  | ΔES (Post-Pre) | 0.12 | 0.01 | 0.08 |  |  |
| O<sub>2</sub> debt (L) | Pre | 17898±6978 | 17031±7956 | 20081±7492 | 0.338 | **0.0096** |
|  | Post | 21139±6820 | 18465±8290 | 20899±9324 |  |  |
|  | ΔES (Post-Pre) | 0.47 | 0.18 | 0.10 |  |  |
| HRR at 1 min | Pre | -20±8 | -19±7 | -17±7 | 0.193 | 0.060 |
|  | Post | -23±9 | -19±9 | -19±6 |  |  |
|  | ΔES (Post-Pre) | 0.35 | 0.05 | 0.30 |  |  |
| O<sub>2</sub> pulse<br>(ml/beat) | Pre | 13,5 ± 3,2 | 13,2 ± 3,5 | 14,4 ± 4,2 | **0.016** | **<0.0001** |
|  | Post | 14,4 ± 3,4 | 14,8 ± 3,2 | 17,1 ± 4,8 |  |  |
|  | ΔES (Post-Pre) | 0,27 | 0,47 | 0,59 |  |  |
|  | p-value Δ<br>(Post-Pre) | **0.024** | **0.033** | **<0.0001** |  |  |

ΔES: effect size, BP, blood pressure; HIIT, high-intensity interval training; HR, heart rate; HRR, heart rate recovery; MICET, moderate-intensity continuous exercise training; RD, recovery
delay; $\dot{V}O_2$ , oxygen uptake. Variables are expressed as means ± SD with time and group x time p-value. Adjusted P-value $\Delta$ (Post-Pre) within group in case of a significant group x time interaction.

## Discussion

In this study, we have assessed the effects of different aerobic exercise training program (Combined HIIT/MICET, LV-HIIT, and MICET) on post-exercise V̇O_2_ kinetics, O_2_ deficit, O_2_ debt, and V̇O_2_ RD in patients with CHD. Our main findings were that: 1) aerobic exercise training improves *τ* V̇O_2_ in particular for higher exercise dose program (Combined HIIT/MICET, MICET); 2) Exercise training did no improves V̇O_2_ RD or other *τ* CPET recovery variables; 3) Exercise training improved O_2_ pulse in particular for higher exercise dose program (Combined HIIT/MICET, MICET). To our knowledge, there is few data on the effect of different exercise training program on post-exercise V̇O_2_ kinetics, V̇O_2_ RD, O_2_ deficit and debt in patients with CHD.

Regarding *τ* V̇O_2_, this was the only CPET *τ* that was improved after exercise training in our CHD patients. We should note that the higher effects size were noted in the combined group (HIIT/MICET) and to a lesser degree in the MICET group (ES:-0.21/-0.40). These two groups had the higher exercise dose. Our results are in agreement with 2 previous exercise training studies in CHF patients. Kemp et al. showed an acceleration of submaximal *τ* V̇O_2_ after 12 weeks of exercise training (HIIT, resistance and respiratory training) in CHF patients ^17^. Similarly, Spee et al showed an improvement of *τ* V̇O_2_ after 12 weeks of HIIT in CHF patients ^18^. In these 2 studies, comparison were done to an inactive control group of CHF patients. This CPET recovery parameter is of great clinical importance, it was shown to be related to lower exercise tolerance, cardiac abnormalities/ischemia and muscle oxidative metabolism in cardiac patients ^8, 12, 13^. These improvements may be explained by several mechanisms that include an increased O_2_ utilization by the peripherals muscles via enhanced mitochondrial function ^30, 31^ and/or and increased O_2_ delivery by the muscle vasculature via an improved vasodilatory capacity ^17, 18^. Another potential mechanism may be due to improved cardiac output during exercise, as suggested by previous studies in CHD patients ^12, 13^, and from our improved O_2_ pulse after training. Regarding other *τ* CPET variables (V̇CO_2_ and V̇E), their kinetics were not improved after training in our patients. Studies on *τ* V̇CO_2_ and *τ* V̇E were mostly realized with CHF patients ^8, 10, 32–34^, and none have explored the impact of exercise training on these variables in cardiac patients. *τ* V̇CO_2_ and *τ* V̇E were shown to be prolonged in the more severe CHF patients vs. less severe ones or healthy subjects ^8, 10, 32–34^. Therefore, *τ* V̇CO_2_ might originate from the CO_2_ production from the aerobic metabolism, the plasmatic buffering of metabolic acidosis by the bicarbonates pool, that should be expired by ventilation. As well, *τ* V̇E value was suggested to reflect the breathing pattern recovery from peak exercise, and stimuli from mecano and chemoreceptors controlling ventilation ^32^.

Regarding V̇O_2_ RD, we showed no impact of exercise training on this variable in our CHD patients. As well, the prevalence of V̇O_2_ RD was not reduced after training for all patients. V̇O_2_ RD is a prognosis marker in CHF patients and is linked to a reduced cardiac output, stroke volume, V̇O_2_ /work slope during exercise and cardiac filling pressure in CHF patients ^6, 11^. This is the first study reporting V̇O_2_ RD value in CHD patients and the impact of different exercise training program on this index. As expected, our V̇O_2_ RD values (from 10.8 s to 14.7 s) are much shorter than previously published in patients with CHF ^6, 11^. As well, this remain unclear how exercise training may impact (shorten or reduce prevalence) this new CPET index. We showed that O_2_ pulse (a surrogate marker of stroke volume) was improved in combined and MICET group, but that does not seemed to transfer to reduction of V̇O_2_ RD values or prevalence.

Regarding O_2_ deficit, this element was not improved after training in our CHD patients. High inter-individual variability in O_2_ deficit values were observed that may explain a lack of significant training effect. As well, we calculated this O_2_ deficit during the warm-up period that may differ from previous studies. Our result disagree form a previous study in CHF patients ^35^ showing that continuous aerobic exercise training reduced the O_2_ deficit and the anaerobic contribution during a constant submaximal exercise test. This O_2_ deficit represents the anaerobic metabolism (ATP/phosphocreatine system and anaerobic glycolysis) plus the O_2_ stored in the blood and muscle (hemo/myoglobin) that is used to produce additional ATP (in addition to ATP produced by aerobic metabolism) during incremental exercise and is dependent of the exercise intensity ^26^. This is particular true at the start of exercise and for intensities above first ventilatory threshold, where O_2_ deficit will increase during a ramp type test ^26^.

For the O_2_ debt, the values were increased after training (time effect) in our CHD patients, in particular for the combined group with a higher ES noted. We should mention that the O_2_ debt after an incremental test has been linked to exercise intensity, this debt being higher for more intense level of exercise ^26^. This might be particularly the case, for the combined (HIIT/MICET) and MICET groups, that improved their V̇O_2_peak (data not shown) and therefore reached a higher exercise intensity post-training.

### Limitations

In our study, data of three prospective randomised control trials were pooled for analysis from one single institution and with the participants composed mainly of men. However, a carefully selected and very homogenous population of patients with CHD was part of this study. We used low volume HIIT protocol (passive recovery) in the HIIT group ^22–24^ that is not the most commonly used in clinical research in patients with CHD ^36, 37^. In addition, short to long HIIT protocols were used in the combined HIIT/MICET group ^21^. Therefore, the exercise volume (or dose) differed between the groups, with a higher dose in the combined one. Regarding CPET kinetics calculation, we used an incremental ramp protocol, and we did not performed a constant work rate test that could have produced more comparable results with previous studies.

## Conclusions

We showed that aerobic exercise training with difference training modalities (HIIT, MICET, combined HIIT/MICET) improved *τ* V̇O_2_ in patients with CHD, with greater effects for higher exercise dose program. Exercise training (whatever the modality) did not improved V̇O_2_ RD time of prevalence, as well as *τ* CPET recovery variables in our patients with CHD sample. Our work confirms the benefits of exercise training in *τ* V̇O_2_ in patients with CHD, suggesting a potential better functional status, cardiac function and muscle oxidative metabolism. Further RCT exercise training studies including more women with CHD or patients with others CHD etiology (post-acute MI or post-acute coronary syndrome) are necessary.

## Supporting information

Suppl Mat S1

## Data Availability

All data produced in the present work are contained in the manuscript

## References

1. Kodama S, Saito K, Tanaka S, et al. Cardiorespiratory fitness as a quantitative predictor of all-cause mortality and cardiovascular events in healthy men and women: a meta-analysis. Jama. 2009;301:2024–2035.

2. Myers J, Prakash M, Froelicher V, Do D, Partington S, Atwood JE. Exercise capacity and mortality among men referred for exercise testing. N Engl J Med. 2002;346:793–801.

3. De Schutter A, Kachur S, Lavie CJ, et al. Cardiac rehabilitation fitness changes and subsequent survival. Eur Heart J Qual Care Clin Outcomes. 2018;4:173–179.

4. Mikkelsen N, Cadarso-Suárez C, Lado-Baleato O, et al. Improvement in VO(2peak) predicts readmissions for cardiovascular disease and mortality in patients undergoing cardiac rehabilitation. Eur J Prev Cardiol. 2020;27:811–819.

5. Vanhees L, Fagard R, Thijs L, Amery A. Prognostic value of training-induced change in peak exercise capacity in patients with myocardial infarcts and patients with coronary bypass surgery. Am J Cardiol. 1995;76:1014–1019.

6. Bailey CS, Wooster LT, Buswell M, et al. Post-Exercise Oxygen Uptake Recovery Delay: A Novel Index of Impaired Cardiac Reserve Capacity in Heart Failure. JACC Heart Fail. 2018;6:329–339.

7. Guazzi M. "Recovering" the Recognition for VO(2) Kinetics During Exercise Recovery in Heart Failure: A Good Practice in Need of More Exercise. JACC Heart Fail. 2018;6:340–342.

8. Cohen-Solal A, Laperche T, Morvan D, Geneves M, Caviezel B, Gourgon R. Prolonged kinetics of recovery of oxygen consumption after maximal graded exercise in patients with chronic heart failure. Analysis with gas exchange measurements and NMR spectroscopy. Circulation. 1995;91:2924–2932.

9. Queirós MC, Mendes DE, Ribeiro MA, Mendes M, Rebocho MJ, Seabra-Gomes R. Recovery kinetics of oxygen uptake after cardiopulmonary exercise test and prognosis in patients with left ventricular dysfunction. Rev Port Cardiol. 2002;21:383–398.

10. Pavia L, Myers J, Cesare R. Recovery kinetics of oxygen uptake and heart rate in patients with coronary artery disease and heart failure. Chest. 1999;116:808–813.

11. Kadariya D, Canada JM, Del Buono MG, et al. Peak Oxygen Uptake Recovery Delay After Maximal Exercise in Patients With Heart Failure. J Cardiopulm Rehabil Prev. 2020;40:434–437.

12. Takaki H, Sakuragi S, Nagaya N, et al. Postexercise VO2 "Hump" phenomenon as an indicator for inducible myocardial ischemia in patients with acute anterior myocardial infarction. Int J Cardiol. 2006;111:67–74.

13. Tajima A, Itoh H, Osada N, et al. Oxygen uptake kinetics during and after exercise are useful markers of coronary artery disease in patients with exercise electrocardiography suggesting myocardial ischemia. Circ J. 2009;73:1864–1870.

14. Girandola RN, Katch FI. Effects of physical conditioning on changes in exercise and recovery O2 uptake and efficiency during constant-load ergometer exercise. Med Sci Sports. 1973;5:242–247.

15. Fukuoka Y, Grassi B, Conti M, et al. Early effects of exercise training on on- and off-kinetics in 50-year-old subjects. Pflugers Arch. 2002;443:690–697.

16. Hagberg JM, Hickson RC, Ehsani AA, Holloszy JO. Faster adjustment to and recovery from submaximal exercise in the trained state. J Appl Physiol Respir Environ Exerc Physiol. 1980;48:218–224.

17. Kemps HM, de Vries WR, Schmikli SL, et al. Assessment of the effects of physical training in patients with chronic heart failure: the utility of effort-independent exercise variables. Eur J Appl Physiol. 2010;108:469–476.

18. Spee RF, Niemeijer VM, Wijn PF, Doevendans PA, Kemps HM. Effects of high-intensity interval training on central haemodynamics and skeletal muscle oxygenation during exercise in patients with chronic heart failure. Eur J Prev Cardiol. 2016;23:1943–1952.

19. Kemps HM, Schep G, de Vries WR, et al. Predicting effects of exercise training in patients with heart failure secondary to ischemic or idiopathic dilated cardiomyopathy. Am J Cardiol. 2008;102:1073–1078.

20. Boidin M, Gayda M, Henri C, et al. Effects of interval training on risk markers for arrhythmic death: a randomized controlled trial. Clin Rehabil. 2019;33:1320–1330.

21. Boidin M, Trachsel LD, Nigam A, Juneau M, Tremblay J, Gayda M. Non-linear is not superior to linear aerobic training periodization in coronary heart disease patients. Eur J Prev Cardiol. 2020;27:1691–1698.

22. Trachsel LD, Boidin M, Henri C, et al. Women and men with coronary heart disease respond similarly to different aerobic exercise training modalities: a pooled analysis of prospective randomized trials. Appl Physiol Nutr Metab. 2021;46:417–425.

23. Trachsel LD, David LP, Gayda M, et al. The impact of high-intensity interval training on ventricular remodeling in patients with a recent acute myocardial infarction-A randomized training intervention pilot study. Clin Cardiol. 2019;42:1222–1231.

24. Trachsel LD, Nigam A, Fortier A, Lalongé J, Juneau M, Gayda M. Moderate-intensity continuous exercise is superior to high-intensity interval training in the proportion of VO(2peak) responders after ACS. Rev Esp Cardiol (Engl Ed). 2020;73:725–733.

25. Guazzi M, Adams V, Conraads V, et al. EACPR/AHA Joint Scientific Statement. Clinical recommendations for cardiopulmonary exercise testing data assessment in specific patient populations. Eur Heart J. 2012;33:2917–2927.

26. Ichikawa Y, Maeda T, Takahashi T, et al. Changes in oxygen uptake kinetics after exercise caused by differences in loading pattern and exercise intensity. ESC Heart Fail. 2020;7:1109–1117.

27. Belardinelli R, Barstow TJ, Nguyen P, Wasserman K. Skeletal muscle oxygenation and oxygen uptake kinetics following constant work rate exercise in chronic congestive heart failure. Am J Cardiol. 1997;80:1319–1324.

28. Garzon M, Dupuy O, Bosquet L, et al. Thermoneutral immersion exercise accelerates heart rate recovery: A potential novel training modality. Eur J Sport Sci. 2017;17:310–316.

29. Girault A, Leprêtre PM, Trachsel LD, et al. Determinants of V□+O2peak Changes After Aerobic Training in Coronary Heart Disease Patients. Int J Sports Med. 2024;45:532–542.

30. Cottin Y, Marcer I, Walker P, et al. [Effect of rehabilitation after myocardial infarction on muscular metabolism. Contribution of phosphorus 31 NMR spectroscopy]. Arch Mal Coeur Vaiss. 1994;87:759–765.

31. Cottin Y, Walker P, Rouhier-Marcer I, et al. Relationship between increased peak oxygen uptake and modifications in skeletal muscle metabolism following rehabilitation after myocardial infarction. J Cardiopulm Rehabil. 1996;16:169–174.

32. Shimizu N, Koike A, Koyama Y, Kobayashi K, Marumo F, Hiroe M. Kinetics of pulmonary gas exchange during and while recovering from exercise in patients after anterior myocardial infarction. Jpn Circ J. 1999;63:459–466.

33. Hayashida W, Kumada T, Kohno F, et al. Post-exercise oxygen uptake kinetics in patients with left ventricular dysfunction. Int J Cardiol. 1993;38:63–72.

34. Riley M, Stanford CF, Nicholls DP. Ventilatory and heart rate responses after exercise in chronic cardiac failure. Clin Sci (Lond). 1994;87:231–238.

35. Mezzani A, Grassi B, Jones AM, et al. Speeding of pulmonary VO2 on-kinetics by light-to-moderate-intensity aerobic exercise training in chronic heart failure: clinical and pathophysiological correlates. Int J Cardiol. 2013;167:2189–2195.

36. Gomes-Neto M, Durães AR, Reis H, Neves VR, Martinez BP, Carvalho VO. High-intensity interval training versus moderate-intensity continuous training on exercise capacity and quality of life in patients with coronary artery disease: A systematic review and meta-analysis. Eur J Prev Cardiol. 2017;24:1696–1707.

37. Ballesta García I, Rubio Arias J, Ramos Campo DJ, Martínez González-Moro I, Carrasco Poyatos M. High-intensity Interval Training Dosage for Heart Failure and Coronary Artery Disease Cardiac Rehabilitation. A Systematic Review and Meta-analysis. Rev Esp Cardiol (Engl Ed). 2019;72:233–243.

