## Supplementary material for "Effects of different exercise training program on post-exercise V̇O_2_ kinetics and V̇O_2_ recovery delay in stable patients with coronary heart disease": Suppl Mat S1

**Supplementary File S1**

**High-intensity interval training (HIIT)**

Following a 5 min warm-up at 30 % of peak work load obtained at the CPET, patients performed 2 to 3 sets of 6-10 minutes with repeated bouts of 15 to 30 seconds at 100 % of peak work load alternated by 15 to 30 seconds of passive recovery. The targeted rating of perceived exertion (RPE: 6 to 20) was set at 15 during the HIIT bouts. The sets were separated by a 5 min active recovery phase at 30 % of peak work load. The training session was terminated by a 5 min cool-down phase at 30 % of peak work load (Trachsel et al. 2019, 2020, 2021).

**Moderate-intensity continuous exercise training (MICET)**

Following 5 min warm-up at 30 % of peak work load, patients performed continuous exercise at 60 % of peak work load for 24 minutes. Patients performed 5 minutes of recovery period at 30 % of peak work load in the end of the session (Trachsel et al. 2020, 2021).

**Combined HIIT and MICET**

Combined aerobic exercise program consisted of a combination of HIIT with starting from short to long bout durations (from 15 seconds to 4 minutes) and MICT (from 20 to 60 minutes). The first four weeks included one short intervals HIIT and two short-duration MICT sessions per week. Then, two medium to long intervals HIIT and one medium to long duration MICT sessions per week were performed from weeks 5 to 12 in both groups (Boidin et al. 2020).

**Resistance training program**

Resistance training consisted of 20 minutes of circuit weight training performed with elastic bands and free weight adapted to each patient's capacity. For each muscle group, patients performed 1 set of 15 to 20 repetitions, followed by a 30-second rest period at a target RPE of 15 (Trachsel et al. 2020, 2021, Boidin et al. 2020).
